# Brain fingerprint changes across the menstrual cycle correlate with emotional state

**DOI:** 10.1101/2023.05.21.23290292

**Authors:** Lorenzo Cipriano, Marianna Liparoti, Emahnuel Troisi Lopez, Laura Sarno, Fabio Lucidi, Pierpaolo Sorrentino, Giuseppe Sorrentino

## Abstract

**Background:** Menstrual cycle (MC) is the cyclical phenomenon with the greatest impact on women’s mood and behavior. To date, little is known about the potential mechanism and neuroanatomical correlates of behavioral and emotional fluctuations across the MC. Brain connectome fingerprinting, a recently introduced technique in the field of brain network analysis, represents a valid approach in assessing the subject-specific connectivity and in predicting clinical impairment in several neurological diseases. Nevertheless, its performance, and clinical utility, in healthy individuals has not yet been investigated.

**Methods:** We conducted the Clinical Connectome Fingerprint (CCF) analysis on source-reconstructed magnetoencephalography signals in a cohort of 24 women across the MC.

**Results:** All the parameters of identifiability did not differ according to the MC phases. The peri-ovulatory and mid-luteal phases showed a less stable, more variable over time, brain connectome compared to the early follicular phase. This difference in brain connectome stability (especially in the posterior brain regions) was able to significantly predict self-esteem, wellbeing, and mood.

**Conclusion:** These results confirm the high reliability of the CCF and its independence from the MC phases and, at the same time, provide neuroanatomical correlates of the emotional and mood aspects that change across the MC.

## INTRODUCTION

In all biological organisms, including humans, there are a large number of phenomena that are organized following precise cycles with time scales ranging from hours (or less) to months ^1^. Human behavior, cognition and mood, for example, are strongly affected by the seasons, the days of the week and also the hours of the day. The cyclical phenomenon by far the most important, as it is linked to the conservation and diffusion of the species, is the reproductive cycle in the female, and in particular the menstrual cycle (MC) in the woman.

However, MC has a relevant impact on women’s lives not only for reproduction, but more generally as regards emotional experiences which can lead to manifest pathological conditions. A recent very large-sample study (3.3 million women across 109 different countries) has highlighted how MC is the cyclical phenomenon with the greatest impact on women’s mood and behavior ^2^. Several studies have shown the strong relationship between psychiatric symptoms and MC, as well as an increased prevalence of psychiatric disorders in women, especially in a time relationship with MC. Representative examples of cyclic-dependent manifestations are perimenopausal depression and premenstrual dysphoric disorders (PMDD) that affect approximately 5-8% of women ^3,4^. Milder (yet clinically significant) premenstrual symptoms (PMS) that significantly impact the quality of life ^5^, affect up to 30-40% of women and, most importantly, the prevalence of at least one premenstrual symptom during the woman’s reproductive life may reach up to 90% ^6^.

The large frequency of these MC-related subjective discomforts suggests that research on this topic should not be based on the categorical adherence to the clinical criteria commonly used to identify mood and/or behavioral disturbances, rather on the notion of psychological well-being which entails more than the mere absence of negative mood ^7^. In this regard, the association between MC and self-esteem, often regarded as a subclinical manifestation of depressive mood, has been suggested ^8–10^. However, generally the knowledge about cycle-related emotional fluctuations, without relevant impact on daily living activities, is sparse. For example, the quality of life assessed through tests that explore the sense of individual emotional well-being or self-esteem is poorly explored.

Low self-esteem predicts future unemployment ^11^, difficulties in social relations ^12^, emotional problems, and risk of developing depression, substance abuse, eating disorders ^9^. It was found that self-esteem was lower in the premenstrual period in women with premenstrual syndrome ^13,14^. Hill and Durante found that self-esteem was higher during the late luteal phase ^15^. However, Edmonds et al. failed to find such an association ^16^. Very recently, an impairment in motor performance associated with lower psychological well-being was observed in the follicular and luteal phases, but not in the ovulatory period. In contrast, Villani et al. ^17^ have shown that well-being does not change across the menstrual cycle.

Despite the overwhelming evidence of the effects of MC on behavior and emotional experiences, very little evidence is available on what the potential mechanisms might be. In order to investigate the role of the MC on specific brain regions and the resultant effects on emotional and behavioral aspects, several structural and functional MRI studies have been conducted. In these studies, the areas most commonly described in relationship with MC fluctuation were hippocampus, amygdala, anterior cingulate cortex, insula, prefrontal cortex and inferior parietal lobe (for an extensive review see ^18^). It is not surprising considering that most of these brain regions are core components of two specific brain networks well-known to be implicated in cognitive and affective disorders: the default mode network (DMN) and the salience network (SN) ^19,20^. Variation in volume and/or activation of these regions was related to affective, cognitive and reward processing as well as negative feedback, thus supporting the potential role of hormonal fluctuation in modulating structure, chemistry and function of specific brain networks and justifying the multiple physiological emotional, cognitive and behavioral changes across the different phases of the MC.

A useful approach for verifying the functional fluctuations of the brain network is brain fingerprinting ^21,22^. This approach allows to compare the pattern of individuals’ functional connectomes (FCs) between different conditions. Such an approach has been previously tested in both health and disease, revealing that people suffering from neurological diseases show reduced identifiability with respect to healthy subjects. The reduced identifiability was able in predicting individual clinical features, giving birth to the concept of the Clinical Connectome Fingerprint (CCF) ^23–26^. In healthy conditions, the same methodology may help outlining brain-behavior relations, by highlighting connectivity patterns variation, as a function of physiological modifications.

In the present work it is hypothesized that the structural and functional changes observed (REF) during MC are able to modify the subject’s specific functional connectome so as to lose the characteristics that make the subject identifiable, even in women who do not suffer from clinically evident alterations of the emotional state across MC. We also hypothesize that the possible modification of the fingerprint can still be associated with a reduction in the psychological well-being state which does not entail a clinically evident negative mood.

To accomplish our aims we performed for each woman a brain fingerprinting based on her whole functional connectome ^27–29^. More specifically, we used source-reconstructed magnetoencephalography (MEG) signals in a cohort of twenty-four healthy, naturally cycling women without premenstrual symptoms and with no signs of anxiety and/or depression. We performed two separate recordings for each subject in three phases, (early follicular, peri-ovulatory and mid-luteal) of MC. After filtering the source-reconstructed data in the canonical frequency bands, we used the phase linearity measurement (PLM) ^30^ to estimate the synchronization between regions, obtaining frequency-specific connectomes. Then, we estimated the identifiability rate of the group at each time point, based on the Pearson’s correlations between connectomes. Furthermore, we compared the similarity between each subject’s connectome at a time point with the group’ connectomes at the other time point, thereby obtaining a “cycle-specific fingerprinting” score of identifiability (IC) for each woman.

To test the hypothesis that the IC score was predictive of the emotional condition, we designed a multilinear regression model from the IC score of each subject. Lastly, we studied the nodal strength of each region of interest (ROI) defining the regions with the greatest contribution in emotional feature prediction.

## RESULTS

### Demographic, sex hormonal and emotional features

In table **1** are shown clinical-laboratory characteristics of our sample. The sex hormonal significant differences between MC phases are in line with the well known hormonal trend across the MC. There were no significant differences in subjective emotional features across the MC.

**Table 1.** Demographic and sex hormonal data

| Demographic features | Mean (SD) |
| --- | --- |
| Age (y) | 26.29 (5.10) |
| Education (y) | 16.54 (2.01) |

| Sex hormone blood levels | Early-follicular (T1) | Peri-ovulatory (T2) | Mid-luteal (T3) | T1vsT2, *p.value* | T1vsT3, *p.value* | T2vsT3, *p.value* |
| --- | --- | --- | --- | --- | --- | --- |
| LH (mIU/ml) | 5.38 (2.34) | 15.91 (11.88) | 5.95 (4.04) | <0.001 | ns | <0.001 |
| FSH (mIU/ml) | 7.28 (1.39) | 7.65 (2.99) | 3.88 (1.16) | ns | <0.001 | <0.001 |
| Progesterone (ng/ml) | 0.30 (0.06) | 1.12 (0.81) | 5.75 (2.61) | ns | <0.001 | <0.001 |
| Estradiol (pg/ml) | 33.94 (12.09) | 134.27 (70.62) | 97.42 (39.72) | <0.001 | <0.001 | <0.05 |

| Affective /emotional tests | Early-follicular (T1) | Peri-ovulatory (T2) | Mid-luteal (T3) | T1vsT2, *p.value* | T1vsT3, *p.value* | T2vsT3, *p.value* |
| --- | --- | --- | --- | --- | --- | --- |
| BAI | 7.62 (7.15) | 7.88 (8.11) | 5.92 (4.70) | ns | ns | ns |
| Rosenberg | 22.58 (4.84) | 23.08 (4.84) | 23.83 (4.15) | ns | ns | ns |
| BDI | 3.71 (3.47) | 3.04 (3.16) | 2.88 (2.82) | ns | ns | ns |
| Wellbeing | 401.88 (50.84) | 400.50 (51.73) | 399.00 (45.40) | ns | ns | ns |

Abbreviations/acronyms: FSH=follicle stimulating hormone; LH = luteinizing hormone. Analysis of variance (ANOVA) of sex hormones according to the MC phases; the p.values of post hoc analysis between the MC phases are shown in the last three columns. Abbreviations: ns = not significant.

### Connectome fingerprint

After False Discovery Rate (FDR) correction, no statistically significant differences, among the three time points, in identifiability parameters (i.e., I-self, I-others, I-diff) were found. Additionally, a linear mixed model, with age, education and phase of cycle set as fixed effects and “subject” as random effect did not show any difference in identifiability parameters values (in each canonical frequency band) according to age, education and, above all, cycle phase. This result demonstrates that, at least limited to our experimental setting which included a two-minute interval between test and retest recording, Iself appears to be an identifiability marker not at all dependent on the menstrual cycle.

### Edge-based identifiability

We used the one-way random-effects intra-class correlation coefficient (ICC) to test the edgewise reliability of the individual FCs. The edges with higher ICC values are those that contributed the most to the identifiability. We studied the distribution of SR values extracted from the fingerprint analysis by adding 100 edges per iteration, from the most to the least stable ones, based on ICC values. Our results showed different contributions of the edges in the identifiability according to the MC phase. The women in the T1 phases quickly reached a complete SR (100%) (**Figure 2A**), maintaining a complete SR up to the 500 edges, where a slowly progressive decay in SR started without ever falling below 90%. The T2 and T3 groups (**Figure 2B and 2C**) also reached an almost complete SR (∼98%) with a few ultrastable edges (right from 100 edges) with a rapid and progressive decline in SR up to 4005 edges (SR of ∼70-75%).

**Fig. 1.**
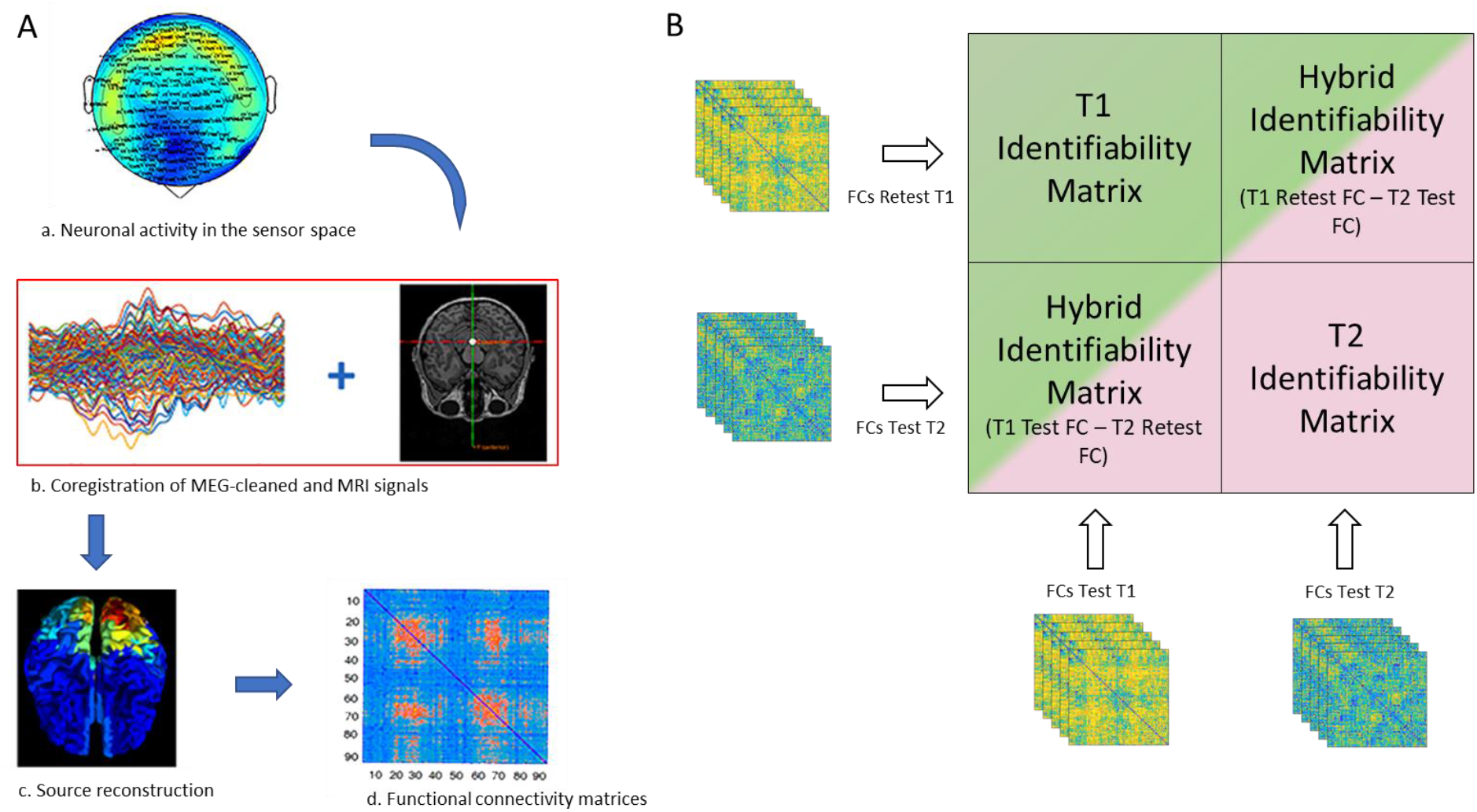
FCs processing and fingerprint analysis. (A) a: the neuronal activity was recorded using a 154-sensors magnetoencephalography (MEG); b: MEG signals cleaned by removing noise and artifacts are coregistrated with MRI scan for source reconstruction (c); d: functional connectivity matrix estimation by PLM. (B) The green and the pink blocks represent the two identifiability matrices of women in two different MC phases (in this explanatory figure: T1 and T2 phases), resulting from the correlation of the test and re-test individual functional connectomes, in each group (according to the MC phase) separately. Hybrid identifiability matrices (IMs) were created by crossing the FCs test of the HC with the FCs retest of the MS and vice-versa. The hybrid IMs return the I-cyclical value of each individual (see methods).

**Fig. 2.**
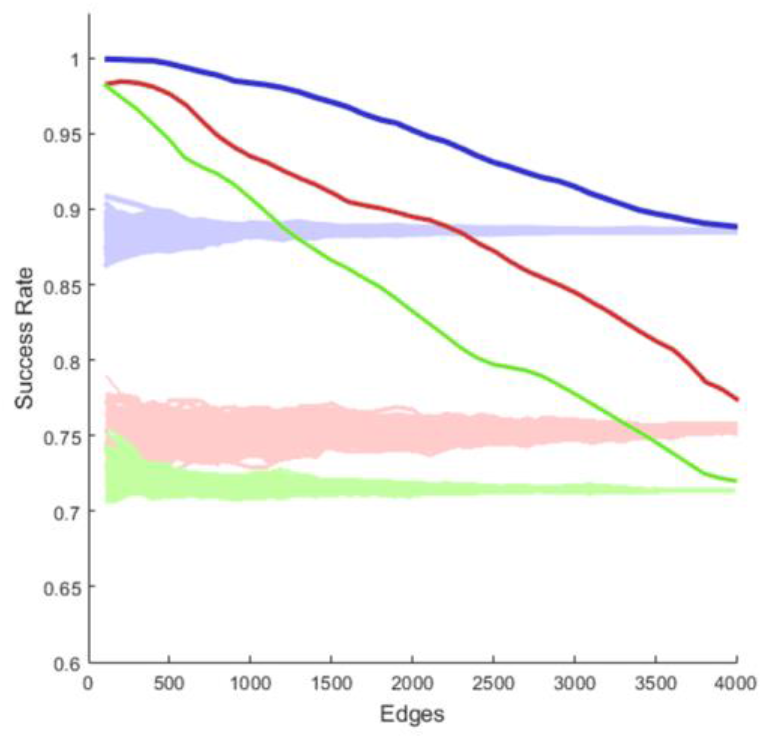
Iterative model of edgewise subject identification. The success rate (SR) distributions of T1, T2 and T3 phases, obtained by adding 100 edges at each step from the most to the least contributing ones, according to the intraclass correlation (ICC) values. The T1 group (blue) quickly reached a complete SR (100%), and a slowly progressive decline but always preserving a great identifiability power. The T2 and T3 groups (red and green, respectively), equally to the T1, quickly reach an almost complete SR but start, after a few hundred edges, an important drop and a progressive loss in subject identification reaching a success rate of ∼70-75% with 4005 edges. The shaded areas of each figure represent the null distributions obtained by adding 100 edges at a time in a random order.

### Multilinear edge-based regression analysis and relationship with subclinical emotional state

We used the IC values to predict emotional conditions assessed by the Rosenberg self-esteem scale, the Ryff’s test (for the six dimensions of well-being), BAI and BDI. We performed different edge-based multilinear regression models using the IC (alongside with age, education level and hormone levels) as a predictor and setting the emotional condition of interest as a dependent variable. The IC with the greatest predictive power was calculated, for each dependent variable, by adding 100 edges in descending order of stability at each iteration up to the full FC.

The IC in the alpha band was significantly able to predict self-esteem level (p-value < 0.01), mood (p-value < 0.01) and almost all the six dimensions of well-being (p-value < 0.01, save the Autonomy). In nearly all the cases (**Figure 3**) the greatest predictive power was achieved when the IC was built with relatively few and highly stable edges (300 for BDI, 200 for Wellbeing and 1200 for self-esteem). IC did not show any significant predictive power on anxiety, assessed by BAI.

**Fig. 3.**
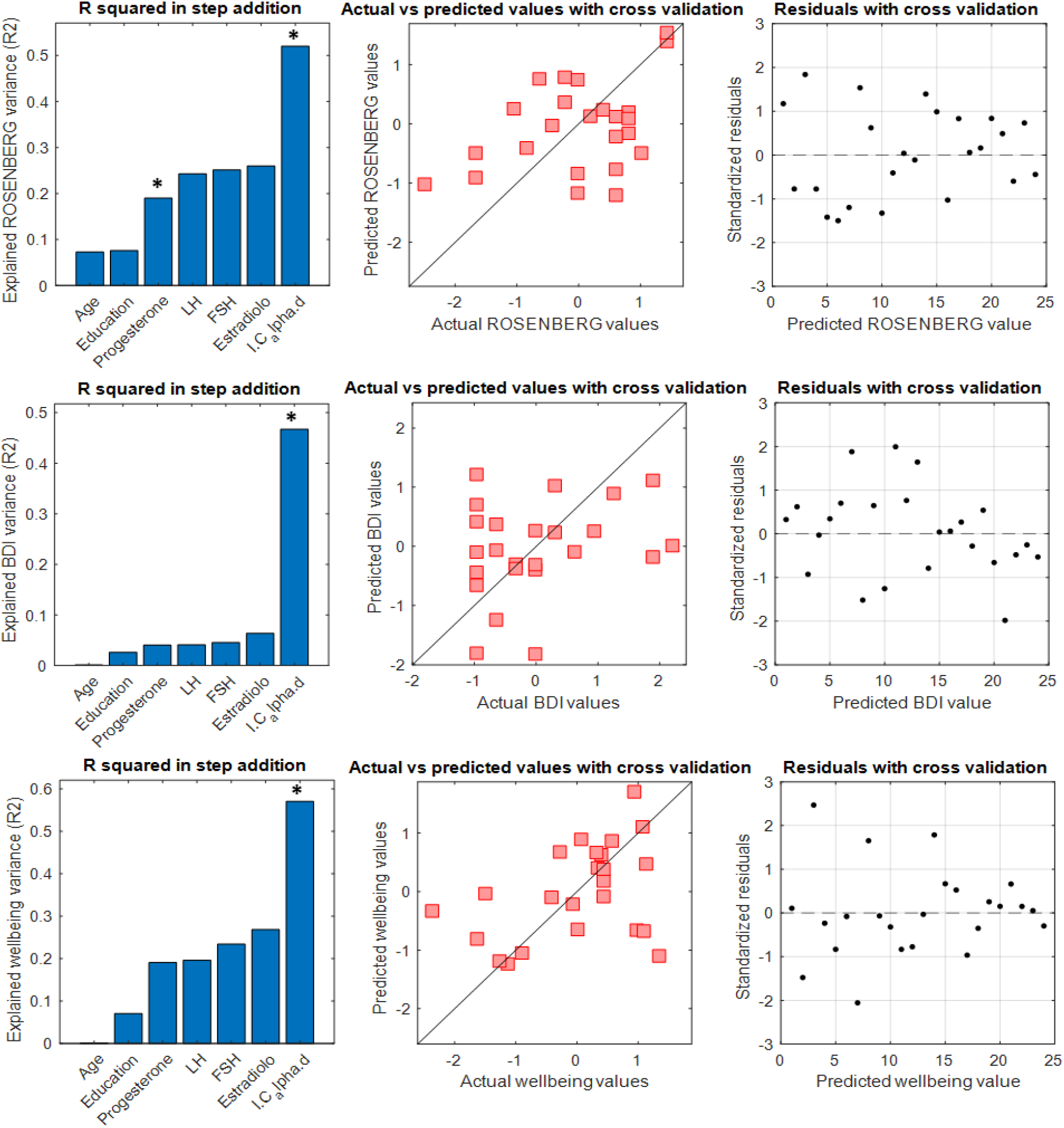
A multilinear regression model with leave-one-out cross validation (LOOCV) was performed to test the capacity of the *I-clinical* to predict self-esteem, depression and well-being in the T2 phase. Panels on the left show the explained variance by the stepwise addition of the six predictors. The significant predictor is indicated with * in bold. The middle panels show the comparison between actual and predicted clinical features. Panels on the right show residuals distribution with cross-validation.

In a Pearson’s correlation, performed for each emotional condition using the IC with greater predictive power, we evaluated the relationship between IC and self-esteem, mood and well-being, respectively. A negative correlation was found between IC and both self-esteem (r = -0.45, p = **0.028, Fig. 4A**) and well-being (r = -0.6, p = **0.002, Fig. 4C**), whereas a positive correlation was observed with BDI (r = 0.62, p = **0.001, Fig. 4C**).

**Fig. 4.**
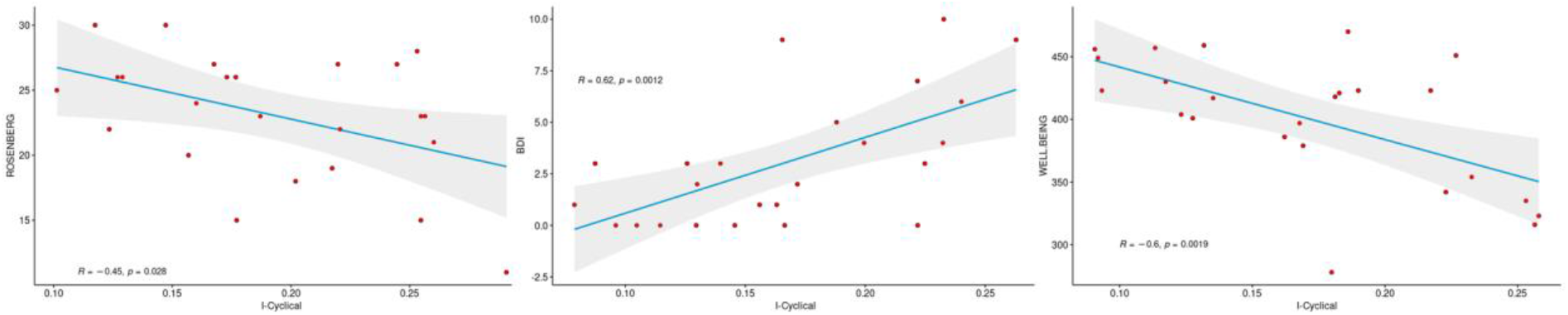
Pearson correlation between cycle-specific fingerprinting and subjective symptoms.

Additionally, we performed the same analyses comparing the other phases of MC between them (i.e. T1-T3 and T3-T2) without finding statistically significant predictive power of IC in the alpha band in predicting the emotional state.

### Regional contribution to identifiability and emotional state

We evaluated the regional contribution of specific brain areas in identifiability and emotional condition. On the basis of the ICC values in the alpha band, we studied the nodal strength of the most reliable edges, thus determining the regions of interest (ROIs) with the greater influence on emotional status and identifiability. We included 1200 edges (because of the best IC values, see above for details) and later we selected the most influential edges (above the 90% percentile). The ROIs with the greatest contribution in identifiability and in predicting emotional outcome were mainly located in the midline and posterior areas such as anterior cingulate, occipital (superior and middle), calcarine, lingual and temporal inferior cortices on the left and parietal superior, occipital (superior, middle and inferior), lingual, cuneus, calcarine and temporal middle cortices on the right (**Figure 5**).

**Figure 5.**
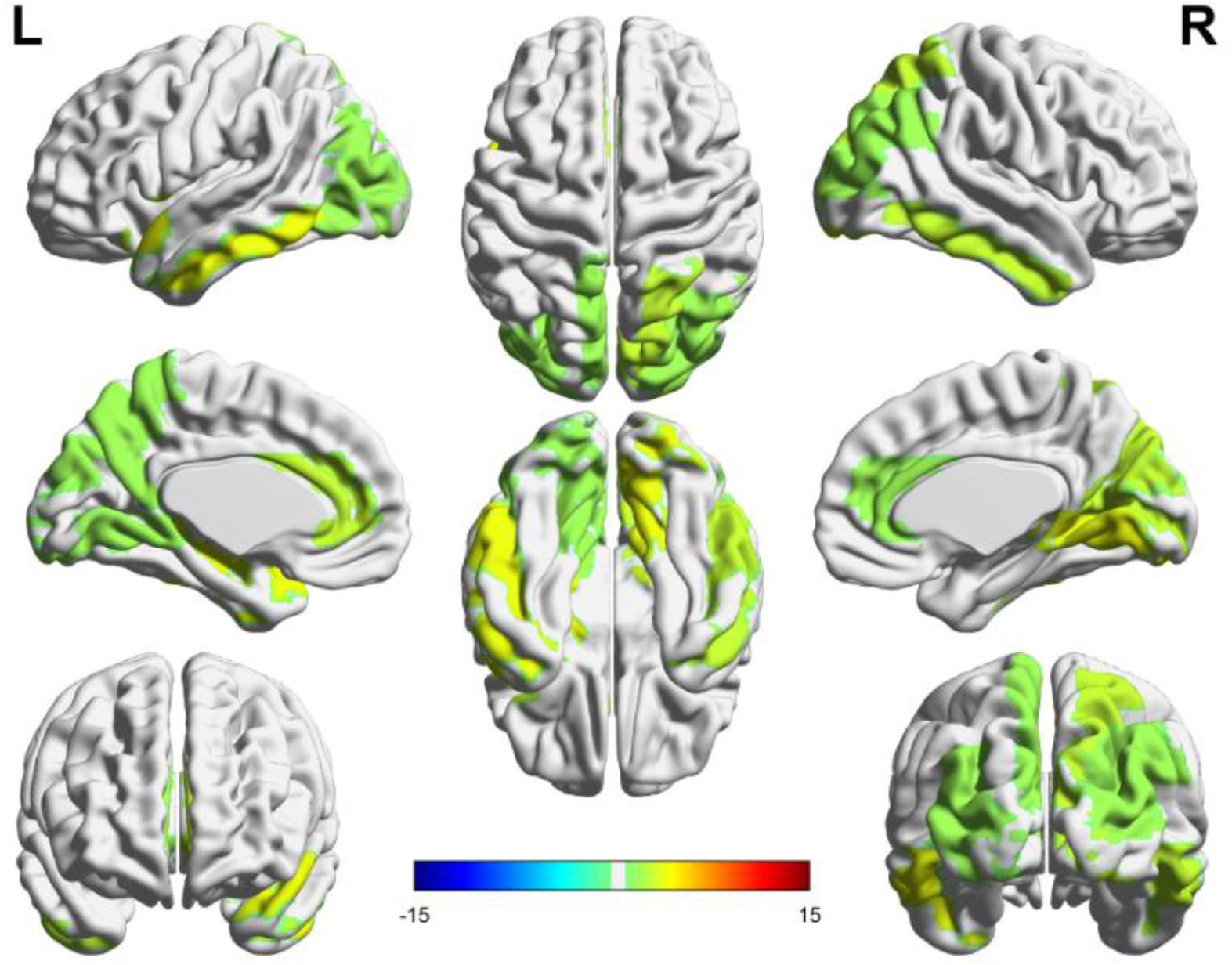
The red areas represent the ROIs with the greatest nodal strength in identifiability and predicting clinical outcome.

## DISCUSSION

In the current paper, we investigated the role that the MC plays in modulating individuals’ functional connectomes (FCs) and how these changes may relate to subjective affective discomforts such as well-being, self-esteem and mood.

We firstly demonstrated that the FC fingerprinting is highly reliable and that, despite the MC affects the brain connectivity, the subject’s specific FCs remain identifiable regardless of the MC phase. Afterward, we investigated the edge contribution in identifiability performing a comparison among early follicular (T1), peri-ovulatory (T2) and mid-luteal (T3) phases. We found that the three groups reached almost a complete SR (100%) with relatively few stable edges. However, the T2 and T3 group identification dropped after only a few hundred edges, descending under 80% of SR with the total number of edges. These data, associated with the low SR of the T2 and T3 null model compared to the T1 suggest a greater disparity of the edge stability in T2 and T3 phase. So, the latter phases seems to have a less stable (i.e. more chaotic, more variable over time) brain connectome compared to T1 phase. This feature was particularly evident in the anterior regions (having a lower number of edges with great identifiability power in the anterior areas since the more stable edges are located mainly in the posterior regions).

These results find partial confirmation in our recent study (unpublished data), where the peri-ovulatory phase was characterized by a greater number of avalanche patterns as compared with the early-follicular phase. The avalanche pattern represents a measure of the brain tendency to explore different configurations, changing quantity and quality of the brain connections in a very restricted time-frame. A higher number of brain reconfigurations, and therefore a greater variability in brain connectivity over time, could explain the difference (also in a wider and averaged time-frame compared to avalanche patterns) in edge stability between T2 and T1 phases. We performed additional analyses to evaluate whether these changes in edge stability between T2 and T1 phases were associated with a sub-clinical worsening of the emotional status. So we built the I-cyclical (IC) on the basis of ICC and performed a multilinear regression model using the IC as a predictor variable of the emotional condition. The IC in the alpha band has been found able to predict self-esteem, well-being and mood.

The reason why the IC with the greatest prediction power was found in the alpha band probably lies in a certain propensity of the MC to influence the alpha rhythm. In fact, previous EEG and MEG studies have shown how the frequency spectrum changes across the MC mostly affecting the alpha band ^31,32^, that is considered strictly dependent on the thalamic activity ^33^. The ovary has been postulated able to influence directly and indirectly (through Meynert nucleus cholinergic pathway) the physiological firing of the thalamus ^34^. In this regard, the interconnection between sex hormones and thalamus has been confirmed by previous neuropathological and neuroimaging studies showing in the thalamic nuclei the presence of enzymes involved in the synthesis of the progesterone (and its neuroactive metabolites) as well as, at functional level (as shown in fMRI studies), a progesterone-associated thalamic activation across the MC (for an extensive review see ^35,36^). Similar evidence is available for estradiol, whose influence on molecular expression in thalamic neurons have been described in over a decade “in vivo” testing ^37^. It is currently hypothesized that sex steroids alter the thalamic GABA pathways ^38^ through the gene expression modulation ^39^ and by influencing the GABA (and GAD) receptor subunits expression ^40^.

To notice that thalamic-modulated alpha rhythm propagation is mainly directed to the posterior brain areas ^41^. This aspect turned out extremely important when we studied which were the regions more connected by the most stable edges in the T2 phase, i.e. the brain areas with the greater contribution in identifiability and in emotional sphere prediction. We found that the brain regions that contributed more to this prediction were mostly located in the posterior areas, thus speculating a modulation of the posterior brain activity operated, at least partially, by sex hormones through a fine regulation of the thalamic firing.

These findings are in line with the previous works both in terms of sex hormone dependent brain areas and self-esteem, well-being and mood neural correlates. With regard to the first point, a follicular/luteal dependence of some brain regions has been demonstrated in fMRI studies that investigated the relationship among sex hormones, ROI-specific brain activity and cognitive/emotional/psychiatric aspects (see ^18,42^). From these studies emerged a relevant MC dependence (usually supported by a positive/negative correlation with sex hormones) of the cingulate cortex (both ACC and the precuneus/PCC system, ^43,44^) and most of the areas reported in our work such as the middle and inferior temporal, lingual and fusiform gyri and parietal and occipital cortex ^43–45^.

The relationship between well-being, self-esteem and mood with posterior and midline brain regions, is in line with the current literature that highlights the importance of said brain areas in self-concept and self-referential aspects as well as in mentalization and motivation circuits.

Concerning self-esteem, the midline structures have been suggested to be involved in the self-referential aspects by encoding self-relatedness of stimuli and by regulating the self-related negative and positive stimuli ^46,47^ whereas the temporal lobe contributes to the self-referential processing and autobiographical memory promoting a memory-based construction of the self ^48,49^. Also posterior brain regions, in particular lingual gyrus, cuneus and the middle occipital gyrus are involved in positive social feedback ^49–51^, contributing to self-referential processing and social cognition ^52–54^ and their activation is associated with negative self-appraisal ^54^, self-criticism ^55^ and the retrieval of self-encoded negative personality traits ^56^.

The midline and posterior areas play an important role also in regulating well-being dimensions. The key role of the ACC in cognitive and emotional features such as conflict monitoring, error detection, motivation, emotion regulation, attention, and cognitive control ^57^ could explain the strong relationship between ACC and well-being. Also, the ACC as well as the PCC, precuneus, superior and middle temporal gyrus are part of the Default Mode Network that have been involved in integrating personally relevant stimuli and maintaining a proper stability in the context of a continuous superfluity of internal and external inputs ^58^. In this way, well-being is commonly seen as the result of the brain’s ability to integrate relevant and significant internal and extrapersonal stimuli in order to safeguard the homeostasis ^59^.

For mood too, the ACC, fusiform and lingual gyri and, in general the occipital regions, have been variably associated with depression risk, antidepressant response and several other aspects related to depression.^60–63^.

Once we evaluated the consistency of our results with the current literature, we investigated the direct relationship between the Icyclical in the alpha band and subjective features. We found a negative relationship between both self-esteem and wellbeing and Icyclical, along with a positive correlation between Icyclical and depression. Since the greater scores at the Ryff and Rosenberg tests are related to positive outcomes whereas higher BDI scores are indicative of worse outcomes, we can interpret these results in a similar way. This means that the higher the stability, across the MC phases, of the connections that involve posterior regions the worse were the outcomes (lower self-esteem and wellbeing as well as increased risk of depression).

In conclusion, in the present study we demonstrate that MC influences the alpha rhythm and that this modulation predominantly affects the posterior regions. Moreover, the reduced stability of functional connections of the posterior areas was associated with higher well-being and self-esteem as well as with better mood. We also showed the ability of a new tool, the Icyclical, to predict subjective emotional conditions, in a significant manner and better than any other predictor (i.e. demographic features and hormonal blood values).

The current work is not free of weakness. First, we did not find (with the exclusion of the Progesterone predictive power for the Rosenberg score) significant relationship between sex hormone blood levels and emotional/brain connectivity parameters, Nevertheless, it is noteworthy to recall that the biochemical changes across the MC are not only about the fluctuation of the four biomarkers investigated in the present article, but include a wider group of other metabolic and endocrine factors. The brain connectome is probably influenced by a complex interaction among all these factors. This could explain why IC (that is itself the result of this complex biochemical interplay) benefits from a greater predictive power as compared to a single hormone. Further studies, including a broader hormone and metabolic investigation, could clarify some of these aspects. Second, we did not find the same high predictive power on the affective/emotional sphere that IC showed in peri-ovulatory phase also in early-follicular and mid-luteal ones. This aspect may be due to a less marked difference between T2-T3 and T3-T1 in edge stability that our small sample size did not allow to unveil. This aspect seems to be supported by the recent study on brain connectivity reconfiguration patterns across the MC (not yet published) that found a clear difference only in T2-T1 comparison.

We think that the current study provides some benefits in the current scientific panorama. Firstly, the functional connectivity change across the MC suggests that in each neuroimaging study protocol (of both MRI and EEG/MEG) we should always take into consideration the MC phase, interpreting critically results that have not been adjusted for the MC phase. Secondly, we confirmed the MC-dependent modulation of posterior brain activity demonstrating that these changes primarily affect the alpha band. We suggest a hypothetical origin of these changes in thalamic regulation operated by sex hormones. This hypothesis will need additional studies for confirmation. Lastly, we demonstrated that our method, i.e. the Icyclical, is able to predict emotional outcomes.

## METHODS

### Participants and experimental protocol

Twenty-four right-handed, native Italian speakers, heterosexual women with regular MC were recruited. Exclusion criteria were: 1) use of hormonal contraceptives (or other hormone regulating medicaments) during the last 6 months before the recording; 2) pregnancy in the last 12 months; 3) chronic use of drugs able to affect the central nervous system; 4) alcohol, tobacco, and/ or coffee consumption 48 hours prior to the MEG recordings; 5) absence of history of neuropsychiatric diseases and premenstrual dysphoric/depressive symptoms. Mood and/or anxiety symptoms were investigated by means of the Beck Depression Inventory (BDI) ^64^ and Beck Anxiety Inventory (BAI) ^65^, using a cut-off below 10 and 21, respectively. To control for the influence of circadian rhythm, the time of testing varied no more than 2 hr among testing sessions. The subjects’ characteristics are shown in Table 1.

All women were tested in three different time points of the MC, that is, in the early follicular phase (cycle day 1– 4, low estradiol and progesterone, T1), during the peri-ovulatory phase (cycle day 13– 15, high estradiol and low progesterone, T2) and in the mid-luteal phase (cycle day 21– 23, high estradiol and progesterone, T3).

At each of the three time points along the cycle all subjects underwent the following examinations: MEG recording, blood sampling for the hormone assay, and psychological evaluation. Hormone assays and ultrasound examination have been performed according to Liparoti et al. 2021 ^66^.

All participants signed a written consent form. All the procedures strictly adhered to the guidelines outlined in the Declaration of Helsinki, IV edition. The study protocol was approved by the local ethic committee (University of Naples Federico II; protocol n. 223/20).

### Psychological evaluation

The psychological assessment was performed at each of the three phases of the MC. To quantify the self-esteem level, the Rosenberg self-esteem scale ^67^ was adopted. The Ryff’s test was administered to examine the six dimensions of well-being (autonomy, environmental mastery, personal growth, positive relations with others, purpose in life, and self-acceptance) ^68^. Finally, in addition to BAI ^65^ and BDI ^64^ tests administered at the first experimental session (as inclusion/exclusion criteria, see above), the tests were re-administered at each time point to exclude the appearance of depressive/anxious symptoms.

### MEG acquisition, preprocessing and source reconstruction

MEG acquisition, preprocessing, source reconstruction and synchrony estimation have been performed according to our previous works ^23,66,69^.

### Fingerprint analysis

At each of the three time points of the MC, we performed two MEG recordings separated by ∼2 minutes, hence calculating two FCs, named test and retest. Based on them (Figure 1), we estimated the brain-fingerprinting of each subject in the three different MC phases. We started by creating frequency-specific identifiability matrices (IM), using the Pearson’s correlation coefficient between the test and re-test FCs. Specifically, the test FC of each subject was correlated to the retest FCs of all the subjects within the same phase of the MC (including her/himself) ^27^. So, the resultant IM embodies, in the main diagonal, the information inherent to homo-similarity (I-self, the similarity between FCs of the same individual), whilst data about hetero-similarity (I-others, i.e., the similarity of that subject’s FC with the whole group) are represented by the off-diagonal elements. Then, we extracted the differential identifiability (I-diff), a score that estimates the subject-specific fingerprint level of a specific brain dataset, by subtracting the I-others value from the I-self value ^27^. Moreover, to define the probability of correctly identifying a specific individual, we calculated the success rate (SR) of subject recognition within a specific group. The SR was computed on the number of times (expressed as percentage) that each subject showed an I-self higher than the I-others (i.e., how many times an individual was more similar to themselves than to another individual of the same group).

Finally, we set out to measure how much each subject’s FC was similar to the mean of the other subject’s FCs in the previous of the MC, by computing the “cycle-specific fingerprinting” score (I-cyclical, IC). Similarly to Sorrentino et al., ^25^ we built two block identifiability matrices by crossing, for each individual, the FC test and re-test respectively. Specifically, the test and retest FCs of each individual were compared to the retest and test FCs, respectively, of each other individual in the previous MC phase (i.e., T2-T1, T3-T2, T1-T3) (Fig. 1). Then, the correlation coefficients were averaged for each individual, obtaining subject-specific IC scores for each MC phase, that represent the similarity to the previous MC phase and as consequence, how much the FC of each individual changed across the MC.

### Edges of interest for fingerprint

To estimate the edgewise reliability of individual connectomes across the test and the re-test recordings, we used the intra-class correlation coefficient (ICC) ^70^, a measure that quantifies the similarity of the elements belonging to the same group. In our case, the higher the ICC values, the greater the stability of an edge over different recordings ^25^. We hypothesized that, in a functional connectome, the edges with higher ICC were the most stable ones over time and then, those that could contribute more to subject identifiability. So, we conducted a fingerprint analysis by sequentially adding them on the basis of their ICC values. We carried out this analysis by adding 100 edges at each iteration (and computed the SR values) starting from the highest ICC value to the lowest. A null model was built by adding the edges in random order, 100 times at each iteration, to validate our findings.

### Fingerprint clinical prediction

Since the IC score describes the similarity of a subject to the subjects in a previousMC phase, it also entails information about how much a given subject’s FC is changed. Following this line of reasoning, we hypothesized that such a parameter may be related to emotional and behavioral aspects that vary in function on the MC. Hence, we built a multilinear regression model to predict self-esteem, well-being and mood scores based on the IC scores and six other predictors: age, education and hormone blood levels (progesterone, estradiol, LH and FSH) ^71^. Multicollinearity was assessed through the variance inflation factor (VIF) ^72,73^. We validated our model using the leave-one-out cross validation (LOOCV) framework ^74^. Furthermore, similarly to the edgewise identification, the multilinear analysis was performed in an iterative scenario where the IC score was calculated using different subset of edges, based on their ICC values. In particular, we calculated the IC by adding 100 edges at each iteration in descending order of stability (from the highest to lowest ICC).

### Statistics

Statistical analysis was performed in MATLAB 2021b. We analyzed all the comparisons among the I-self, I-others and I-diff values using permutation testing, where the labels of the two groups were randomly allocated 10000 times. Thus, we obtained a null distribution of the randomly determined differences by computing the absolute value of the difference of the group averages at each iteration ^75^. The relationship between variables was studied with Pearson’s correlation and the results were corrected for multiple comparisons using the False Discovery Rate (FDR) ^76^, setting the significance level at p-value < 0.05.

## Data Availability

The data that support the findings of this study are available from the corresponding author,
GS, upon reasonable request

## Statements and Declarations

### Funding

Ministero Sviluppo Economico; Contratto di sviluppo industriale “Farmaceutica e Diagnostica” (CDS 000606); European Union “NextGenerationEU”, (Investimento 3.1.M4. C2) of PNRR

All authors declare that they have NO affiliations with or involvement in any organization or entity with any financial interest or non-financial interest in the subject matter or materials discussed in this manuscript.

This study was performed in line with the principles of the Declaration of Helsinki. The study protocol was approved by the Local Ethics Committee (University of Naples Federico II; protocol n. 223/20).

Written informed consent for publication was obtained from all participants.

## Declarations

### Author contributions

Concept and design: LC, ETL, PS, ML. Data collection, analysis and interpretation: ML,LS ETL, FL, LC. Drafting of the manuscript: LC and GS. Critical revision of the manuscript for important intellectual content: FL, GS, LS.

### Data availability

The data that support the findings of this study are available from the corresponding author, GS, upon reasonable request.

### Ethical statement

All procedures performed were in accordance with the ethical standards of the institutional research committee and with the ethical standards laid down in the 1964 Declaration of Helsinki and its later amendments.

### Informed consent and consent to participate

Written informed consent has been obtained from all participants.

